# Adolescent Anxiety and TikTok. An exploratory study

**DOI:** 10.1101/2022.09.27.22280435

**Authors:** Andrey Zheluk, Judith Anderson, Sarah Dineen – Griffin

## Abstract

**INTRODUCTION:** Social media is ubiquitous in adolescents’ lives. TikTok is a medium primarily used by adolescents and young adults under 30 years. TikTok is thus an appropriate social media platform with which to examine discussions of anxiety among this age cohort. In this exploratory mixed-methods study we aimed to evaluate the scope of anxiety content available on TikTok in English in December 2021, and to further develop methods for analysing TikTok content.

**METHOD:** We analysed a data set of 147 TikToks with the hashtag #anxiety. The data set consisted both of metadata and TikTok videos. This data set represented 18% of all TikToks featuring the hashtag #anxiety in December 2021. We examined the following research questions (RQs). RQ1: What are the creator identities reflected in the final data set in this study?; RQ2: What are the metadata characteristics of the TikToks in the final data set?; RQ3: What are the anxiety content themes in the final data set?; and RQ4: What are the characteristics of the data set based on an anxiety management reference checklist?

**RESULTS:** Influencers were the most frequent creator identity in our data set. Influencers comprised 85.5% of the 147 TikToks in our final data set. We coded 79 (54%) female and 45 male (31%) influencers. We found male influencers created the most played (mean 81 147 062), and most liked (mean 1 510 585) TikToks. We found content themes varied by influencer gender. Notable finding were: the greater use of humour by males (22.7% males; n=10, and females 12.7%; n=10); and inspiration (38.6%; males n=17; and 13.9%; females n=11). Among female influencers, we identified self-disclosure as the most common theme (n= 40 and 50.6% compared with n=11 and 25% male influencers). Overall, we found limited references to evidence-based anxiety self-care content in our final data set.

**DISCUSSION:** We suggest that the TikToks in our data set were primarily directed at raising awareness of and de-stigmatising anxiety symptoms. TikTok anxiety content may be viewed by adolescents for emotional self-regulation, beyond evidence-based health information seeking. Self-disclosure on TikTok may also provide symptomatic relief to adolescents with anxiety. We suggest that gender is a salient consideration when considering TikTok content.

**CONCLUSIONS:** Our content findings are broadly consistent with previous research into adolescent use of social media. This research also provides methodological insights for researchers and clinicians seeking to understand TikTok, and to develop engaging content targeted at the specific concerns and preferences of adolescent TikTok consumers.

## INTRODUCTION

Social media is ubiquitous in adolescents’ lives. Through conducting this exploratory mixed-methods study, we aimed to evaluate the scope of anxiety content available on the social media platform TikTok in December 2021, and to further develop methods for analysing TikTok content. TikTok is a medium primarily used by adolescents and young adults under 30 years. TikTok is thus an appropriate social media platform with which to examine discussions of anxiety among this age cohort. This examination of the characteristics of popular TikTok videos may assist researchers and clinicians in understanding this rapidly expanding medium and to develop content that may more effectively engage with young people experiencing anxiety.

### Epidemiology of anxiety

Anxiety disorders represent the second leading cause of disability among all psychiatric disorders. Anxiety disorders are associated with psychiatric and medical comorbidities and role impairments in occupational and social domains [1]. Up to 34% of individuals report experiencing an anxiety disorder over the course of their life, with females twice as likely to experience anxiety as males [2].

The incidence of anxiety is highest during adolescence and early adulthood [3]. We have defined adolescence as the ages between 10 and 24 years [4]. Among adolescents, anxiety disorders are the most common class of mental disorders, with a 12-month prevalence rate of 24.9% [5]. In addition to a high prevalence of anxiety, adolescents experience profound biological, psychological, and social development during this age span.

### Clinical features and treatment of anxiety

Anxiety covers disorders including Generalised Anxiety Disorder (GAD), PTSD, panic disorder, and social anxiety [6]. Clinical features of anxiety include excessive and difficult to control worry, fatigue and poor sleep patterns, poor concentration, muscle tension, and impaired social functioning. Physical symptoms can include shortness of breath, a pounding heart and trembling hands [7,8].

Anxiety disorders can be treated successfully with modifications to lifestyle, medication and psychological therapies [7,9]. However, anxiety disorders are underdiagnosed and underreported. Studies have suggested that between 20 and 50% of anxiety cases are formally diagnosed [10]. Of those formally diagnosed, and a smaller proportion receiving a combination of medication and psychotherapy [11].

### Social media and mental health

Researchers have offered mixed opinions on the relationship between social media use and mental health. Social media are internet-based communication platforms that allow users to interact, share information and create web content in online communities [12]. In 2021 the most popular social media across all age groups were YouTube and Facebook [13]. In 2021 TikTok was the most downloaded mobile phone app globally [14], and the most rapidly growing social media channel [15].

TikTok is most popular among adolescents. In 2020, 69% of the TikTok’s user base were aged 13 to 24 years [16]. The COVID-19 pandemic coincided with a rapid increase in TikTok use by adolescents across the globe. During 2020 social media use increased by 61% [17]. Between February and April 2020, U.S. children aged between 4 and15 years spent 13 % more time on YouTube, 16 % more time on TikTok, and 31 % more time on Roblox [18]. A 2022 Pew Research study found YouTube is the most popular social media used by 95% of teens, followed by 67% for TikTok [19]. Pew research further reported the gendered character of social media platforms. Adolescent males are more likely to say they use YouTube, whereas females are more likely to use TikTok and Instagram. Pew Research further reported that 97% of teens use the Internet daily, with 95% using smartphones. In 2022, Pew found 46% of teens reported that they were online constantly, compared with 24% in 2015 [20]. Social media use among adolescents in 2022 is ubiquitous.

The ubiquity of social media use by adolescents has attracted increasing scholarly interest. Researchers have analysed social media use from perspectives including social influence theory [21], social presence theory [22] and motivation theory [18]. Uses and gratification theory has been among the most influential theoretical approach to analysis of social media content [23].Researchers have deployed uses and gratifications theory to adolescent social media use. From this theoretical perspective adolescent TikTok users satisfy both their cognitive and affective needs [24–26].

Researchers have examined the positive and negative associations between mental health and social media use by adolescents. The reported positive benefits of social media include identity development and maintaining social connections [27,28]. Other investigators have suggested that TikTok creators offer health advice or discuss health-related topics that receive significant engagement [29,30].

Researchers have also reported increased anxiety associated with social media use. Some researchers have suggested that increased use of social media by adolescents is linked to increased risk of anxiety disorder [31–34 35]. Other reported problems include social comparison, disordered eating; substance use; self-harm and depression [29,36]. In summary, scholarly opinion remains divided on the harms and benefits of social media use by adolescents.

### COVID-19 and anxiety

COVID-19 may have contributed to both anxiety disorders among adults and adolescents. Galea and colleagues suggested COVID-19 likely produced anxiety and depression at population scale similar to earlier crises such as September 11 in the United States [37]. A 2021 systematic review of mental health peer reviewed literature published during COVID-19 found researchers reporting increased anxiety among half of the population across all ages [38]. During COVID-19 adults were affected by social isolation, fear of the infection and financial impact [39]. Most people in COVID-19 social isolation had no access to mental healthcare [40]. During COVID-19, many individuals were forced instead to rely on themselves, using self-help, self-medication and self-care.

The COVID-19 pandemic during 2020-22 increased anxiety among adolescents. Investigators have suggested that the emotional stresses during the pandemic may translate to longer term mental health problems for adolescents as they transition to adulthood [41]. Other researchers have reported that the adolescent use of social media during COVID-19 may have played a positive role. Social media and additional time with family may have provided adolescents with social connection [28,42,43]. Further, social media may also have assisted adolescents with managing their mental health, through the disseminating health-related recommendations, psychological first aid [44], and emotional, and peer support [45,46].

### Researching social media and anxiety

Social media are a novel and publicly accessible record of adolescent behaviour. Through social media interactions, users describe their activities, ideas and emotions [47]. While describing these experiences social media users also create a record of their social behaviours. It is the creation of what is effectively a publicly accessible database of adolescent behaviours that has attracted the interest of population health researchers and mental health clinicians [48]. As TikTok use has increased, so has scholarly interest in the medium. Researchers have suggested that TikTok offers a creative space for adolescents to explore social worlds and to develop their identity [43,49]. Mental health researchers have described TikTok as “the ideal platform to disseminate public health information” to adolescents [43,49], and “a new playground for the child psychiatrist” [50].In summary, TikTok providers a rich source of data to research anxiety behaviours among adolescents.

## METHODS

In this exploratory mixed-methods study we aimed to evaluate the scope of anxiety content available on TikTok in English in December 2021, and to further develop methods for analysing TikTok content. We examined the following research questions (RQs). RQ1: What are the creator identities reflected in the final data set in this study?; RQ2: What are the metadata characteristics of the TikToks in the final data set?; RQ3: What are the anxiety content themes in the final data set?; and RQ4: What are the characteristics of the data set based on an anxiety management reference checklist?.

This study is based on methods of social media analysis developed by Zheluk, Anderson and Dineen Griffin for their analysis of TikTok low back pain content [30]. We adapted the methods to this study in the following ways. First, we have assumed that anxiety TikToks are viewed by for self-care guidance only (i.e. TikToks are viewed independent of health advice). Second, we have used a broad definition of anxiety. All TikToks including the hashtag anxiety were included in our data set. As a consequence, anxiety may be the primary theme, or secondary to another physical or mental health problem described by the TikTok creator. Third, we did not examine information quality. Information quality instruments have been widely used to evaluate online content [51,52]. Several researchers have suggested that information quality instruments such as DISCERN and JAMA may not be suitable for analysis of social media video content.

### Data collection and cleansing to obtain the final data set

We first identified anxiety - related TikTok videos for analysis through three steps.

### Step 1: Selection of search terms

TikToks are presented to viewers through either an algorithm, or via hashtag (#) based search. Algorithmic presentation of TikToks is automated and based on a user’s previous viewing history. Hashtags by contrast, allow discovery of specific themed content on TikTok. We examined alternative search terms including stress, panic, fear and distress. We identified the hashtag “#anxiety” as the most viewed relevant English - language TikTok search term. See table 1 related terms.

**TABLE 1:**
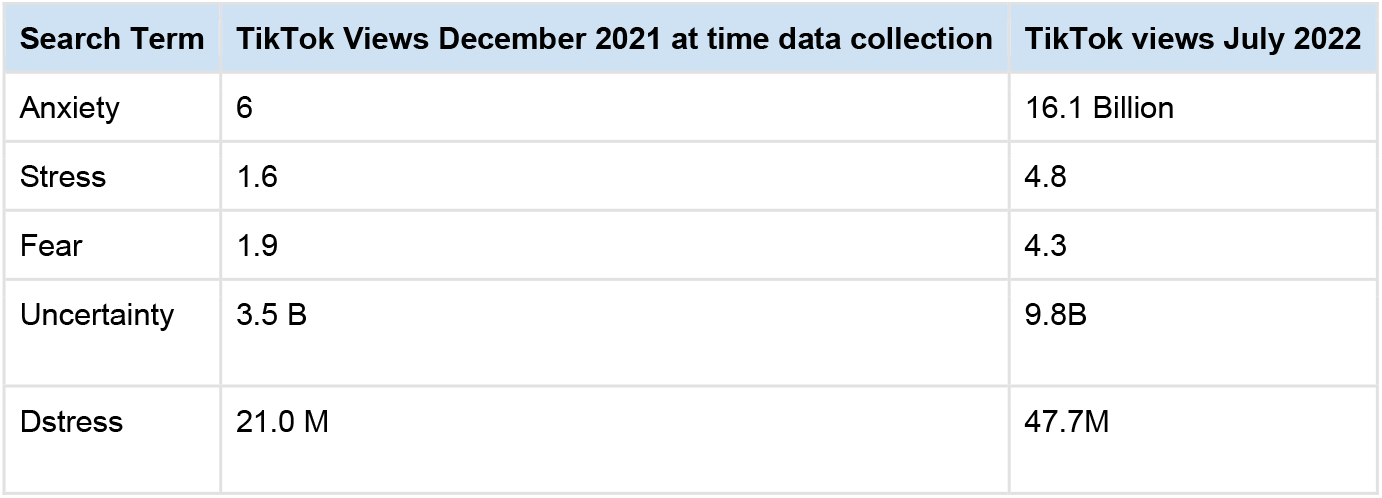
Search terms related to anxiety.

### Step 2: Raw Data Set (stress, anxiety, panic, fear, distress)

We initially identified a raw data set of the 200 most viewed TikTok videos by searching for #anxiety in the TikTok app on December 12, 2021. These 200 TikTok videos in our raw data set had been viewed 6 billion times. This number of views represents the total global views of each individual TikTok tagged with #anxiety since the time of publication. Our raw data set represented approximately 23% of all TikTok views tagged with #anxiety by December 2021 [36]. Second, we downloaded the metadata for these TikTok videos. We used the TikTok scraper and downloader tool (TSDT) version 1.4.36 in order to scrape the metadata for #anxiety TikToks to produce the raw data set [53]. The TSDT allows for downloading of metadata and video content for a specified number of videos for a specific TikTok hashtag. The relevant fields contained in the metadata include the number of views of each video at the specified date, length, internet address, publisher, and date of publication.

### Step 3: Cleansing the Raw Data to Produce the Final Data Set

Our raw data set consisted of 200 TikToks. We excluded 53 non-English language, duplicate, deleted, and TikToks not relevant to anxiety as a clinical condition. The final data set thus consisted of 147 TikTok videos in English that were relevant to anxiety. Relevant videos were those that featured anxiety as the primary theme, or secondary to another condition or life circumstances, and had not been deleted by the TikTok creator between data collection and analysis. The final data set represented approximately 18% of all TikTok views tagged with #anxiety globally in December 2021. [See Appendix 1 final data set].

### Research Questions

We used the final data set in order to answer four RQs.

### RQ1: What are the TikTok Creator Identities in the Final Data Set?

We aggregated TikToks by creator identity in order to segment and categorise the content in our final dataset. We define TikTok creators as the owners of accounts. TikTok creators may not always appear in videos but are generally identifiable by gender. In the mental health domain, researchers have suggested that the highest proportion of the most popular YouTube videos about GAD [47] and depression [54] are created by consumers.

Researchers have suggested that aggregated creator identities provide insights into content themes. Zheluk and Maddock demonstrated differences in volume of YouTube views, and concordance with scientific evidence based on creator identities [52]. In their research, physician created videos were in aggregate most closely concordant with scientific evidence. In another study, Zheluk and colleagues found chiropractors consistently produced TikToks with limited scientific content [30]. These limited scientific content chiropractic TikToks were consistently the most viewed.

We used a general inductive approach to identify TikTok creator identities in the final data set. Thomas describes a general inductive approach as an appropriate method for developing an analytic framework out of raw data linked to the aims of a research project [55]. See Table 2 for creator identity definitions. We identified the following creator identities: male influencer; female influencer; medical clinician; non-medical clinician and animations identity.

**TABLE 2.** Creator identity definitions.

| Creator identity | Views December 2021 |
| --- | --- |
| Male influencer | Tik Tok created by a non - clinician of male appearance. This code is independent of the number of views of a TikTok, and may include TikToks created by individuals living with anxiety. |
| Female influencer | Tik Tok created by a non -clinician of female appearance. This code is independent of the number of views of a TikTok, and may include TikToks created by individuals living with anxiety. |
| Non- medical clinician | TikTok was created by a self-identified psychologist, counsellor, social worker, coach, youth worker or other self-identified provider of mental health services. |
| Medical clinician | TikTok was created by a self-identified medical practitioner. |
| Animated TikToks | Tik Tok content is animated. TikTok created by an individual of unspecified gender. Creator gender and affiliation is not evident from TikTok content, or the creator's TikTok identity. |

“Influencer” was overwhelmingly the most prevalent creator identity in our final data set. We defined influencers as consumers or otherwise non-organisationally affiliated TikTok creators, irrespective of the number of followers. We divided creator identities by gender. In addition, we described a creator identity “animations”. In the “animations” identity, influencer gender or professional identity could not be identified. We used aggregated creator identities to investigate each of the four research questions. For example, we examined all TikToks coded in the “influencer” category to analyse the metadata, the anxiety content themes, and concordance with the reference checklist in the final data set.

### RQ2: What are the metadata characteristics of TikToks in the final data set?

Researchers have incorporated the analysis of metadata elements including views, likes, shares, length when analysing mental health and social media use [54,56]. Researchers have associated aggregated creator identities with metadata elements for both TikToks and YouTube [30,52].

In this study we examined the metadata characteristics of the aggregated TikToks in each creator identity. We used the aggregated creator identities as the units of analysis to examine TikTok metadata. We used the metadata obtained via the TSDT to answer this question. In this study, we used the following data fields: days since published to December 12, 2021, views, video duration, likes, and shares. Through this approach, we were able to describe the metadata characteristics of the final data set.

### RQ3: What are the anxiety content themes in the final data set?

We first identified common anxiety content themes through inductive thematic analysis. These themes were humour, inspiration, self-disclosure, ASMR, “I get overwhelmed” song trend and “identical twins” trend. These themes were used as codes to analysis the final data set. See Table 3. We combined apriori themes with high frequency patterns identified in our final data set. First, we developed a codebook to ensure consistency of thematic coding and used coder consensus to ensure reliability. Second, we then used the aggregated creator identities (eg male influencer, female influencer) as the unit of analysis to examine the content themes in the final data set.

**TABLE 3:** Anxiety content themes codebook.

| Theme | Definitions |
| --- | --- |
| Humour | TikTok is primarily focused on humour and associated with anxiety<br>OPTIONS: yes/no |
| Inspiration | TikTok is from a third person perspective and provides education and /or inspiration to individuals affected by anxiety. Includes religious inspiration<br>OPTIONS: yes/no |
| Self-disclosure | TikTok is from a first person perspective and describes the experience of living with anxiety by an individual affected by anxiety<br>Self-disclosure involves intentionally sharing personal information about oneself. OPTIONS: yes/no |
| ASMR | Video music, words or images that are primarily directed at soothing symptoms of anxiety (ie viewer is not required to perform activities to soothe anxiety symptoms eg Autonomous Sensory Meridian Response (ASMR)<br>OPTIONS: yes/no |
| “I get overwhelmed” song trend | The use of audio from the song “I get overwhelmed” by US musicians Royal and the Serpent song<br>OPTIONS: yes/no |
| “Identical twins” trend | The use of TikTok editing to provide the impression that one individual is engaged in a conversation with an identical twin.<br>OPTIONS: yes/no |

We incorporated two TikTok trends as themes through our coding process. TikTok Trends may be music, editing styles, or other features common across content creator categories [57,58]. For example, we have described the “identical twins” trend in the final data set. The identical twins trend refers to use of TikTok editing to provide the impression that one individual is engaged in a conversation with an identical twin. See for example the TikTok by influencer Connor DeWolfe TikTok [59]. We enumerated the use of audio from the song “I get overwhelmed” by US musicians Royal and the Serpent [60] as a second trend in our final dataset. Incorporating trends as themes in analysis highlights a feature of TikTok content that may not be familiar to mental health clinicians and researchers.

We did not include COVID-19 as an anxiety content theme in our final dataset. The data for this study was collected during the Omicron wave of COVID-19. We found only one mention of covid in the text associated with the final data set. The mention of covid was from a male influencer. The mention was single hashtag among multiple hashtags for a single TikTok video (#covid).

### RQ4: What are the characteristics of the data set based on an anxiety management reference checklist?

Researchers have used expert opinion and reference lists as an approach to determining the scientific veracity of social media content [61,62]. For this study we created a reference checklist based on the anxiety self-management items described by In the Royal Australian and New Zealand College of Psychiatrists (RANZCP) guidelines [7]. The checklist included items that an individual may reasonably be expected to independently initiate as part of a self-care intervention for anxiety after viewing a TikTok. See Table 4. The items in the reference checklist used in this study were Cognitive Behavioral Therapy (CBT) treatment; non-CBT psychological treatment; medication use; education; self-monitoring; positive behaviour; peer engagement; and clinician involvement. These categories are similar to categories derived inductively from coding of public presentations of depression in YouTube videos by Devendorf and colleagues [54]. These researchers reported treatment and self-care categories including medication, therapy, diet, exercise, mindful practices, and alternative treatments.

**TABLE 4.** Anxiety management reference checklist codebook.

| **Anxiety self management strategy** | **Description** |
| --- | --- |
| Cognitive Behavioural Therapy (CBT) | TikTok refers to cognitive behavioural therapy.<br>OPTIONS: yes/no |
| Non-CBT psychological therapies | TikTok refers to evidence-based non-CBT non medication treatments for anxiety described in RANZCP guidelines. This includes evidence based complementary therapies, breathing, mindfulness, bibliotherapy)<br>OPTIONS: yes/no |
| Medication use | TikTok refers to use of physician prescribed medication or references to physician prescribed medication.<br>OPTIONS: yes/no |
| Education | TikTok refers to education about prognosis, personal experiences,evidence, symptoms or other dimension of anxiety from a third person perspective<br>OPTIONS: yes/no |
| Self monitoring | Titkok refers to awareness of symptoms and active self-monitoring of anxiety symptoms. Anxiety symptoms may be secondary to another physical or mental health issue. Self monitoring includes awareness of internal state ASMR<br>OPTIONS: yes/no |
| Positive behaviours | Titkok refers to exercise, diet, sleep, and other lifestyle factors<br>OPTIONS: yes/no |
| Peers and social /emotional support | Titkok refers to peer support, friendship. This includes parental support, support groups, and staged same-person TikTok discussion about anxiety related issues<br>OPTIONS: yes/no |
| Clinician involvement | Titkok refers to medical or non medical clinician involvement<br>OPTIONS: yes/no |

### Intercoder reliability

Coding was conducted by the three authors of this study. Intercoder reliability was achieved through intercoder consensus [63]. We reviewed the coding of creator identities, anxiety themes, and an anxiety reference list. Author A initially coded the final data set. Authors B and C subsequently coded the same final data set. We identified an initial discrepancy of 20 individual items within the 147 TikTok videos in the final data set. Following team negotiations, changes to TikTok coding and changes to the codebook were introduced. The final data set and codes represent a team consensus position.

## RESULTS

### RQ1: What are the TikTok Creator Identities in the Final Data Set?

We examined the following author identities in the final dataset: male influencers, female influencers; animations; medical clinicians; and non-medical clinicians. See TikTok creator identities Table 5. Influencers were the most frequent creator identity in our data set. Influencers comprised 85.5% of the 147 TikToks in our final data set. We coded 79 (54%) female and 45 male (31%) influencers out of a total of 147 TikToks in our final data set. Medical clinicians were the smallest creator identity. The same physician produced the three TikToks in our final data set.

**TABLE 5.** TikTok creator identifies in the final data set.

| Identity | Number and percentage of total |
| --- | --- |
| Female influencers | 79 (53.7%) |
| Male influencers | 45 (30.6%) |
| Non Medical clinicians | 12 (8.2%) |
| Animations | 8 (5.4%) |
| Medical clinicians | 3 (2%) |
| **TOTAL** | **147 (100%)** |

### RQ2: What are the Metadata Characteristics of Videos in the Final Data Set?

We examined the date of TikTok publication, as well as the number of views, likes and comments for each creator category. See Table 6. We found male influencers created the most played (mean 81 147 062), and most liked (mean 1 510 585) TikToks. Non-medical clinicians created the most shared TikToks (mean 42 398). The most commented upon (mean 22600) and longest (mean 102 seconds) TikToks were produced by physicians. However, the small number of TikToks created by physicians (n=3), and non -medical clinicians (n= 12) suggests these results should be interpreted with caution.

**TABLE 6.** Metadata characteristics of TikToks in the final data set by creator identity.

| Creator identity | Mean duration | Mean likes | Mean shares | Mean views | Mean comments |
| --- | --- | --- | --- | --- | --- |
| Male influencer n=45 | 22 | 1510585 | 33274 | 81147062 | 14787 |
| Female influencer n=79 | 30.5 | 1209851 | 31928 | 6681013 | 15073 |
| Animated<br>n=8 | 21.4 | 1174113 | 36735 | 6525000 | 13256 |
| Non medical clinicians<br>n=12 | 37.7 | 1221045 | 42398 | 5690909 | 19861 |
| Physicians<br>n=3 | 102 | 1013667 | 25700 | 4733333 | 22600 |

### RQ3: What are the anxiety content themes in the final data set?

We examined the following anxiety content themes. These themes were humour, inspiration, self-disclosure, ASMR, “I get overwhelmed” song trend and “identical twins” editing trend. See Table 7. We found content themes varied by influencer gender. Notable finding were: the greater use of humour (22.7% males; n=10, and females 12.7%; n=10); and inspiration themes (38.6% ; males n=17; and 13.9%; females n=11). Among female influencers, we identified self-disclosure as the most common theme (n= 40 and 50.6%) among female influencers, (n=11 and 25% male influencers). We identified 13 TikToks in our final data set that feature “identical twins” trend and 10 TikToks that used of audio from the song “I get overwhelmed” by US musicians Royal and the Serpent [60].

**TABLE 7.**
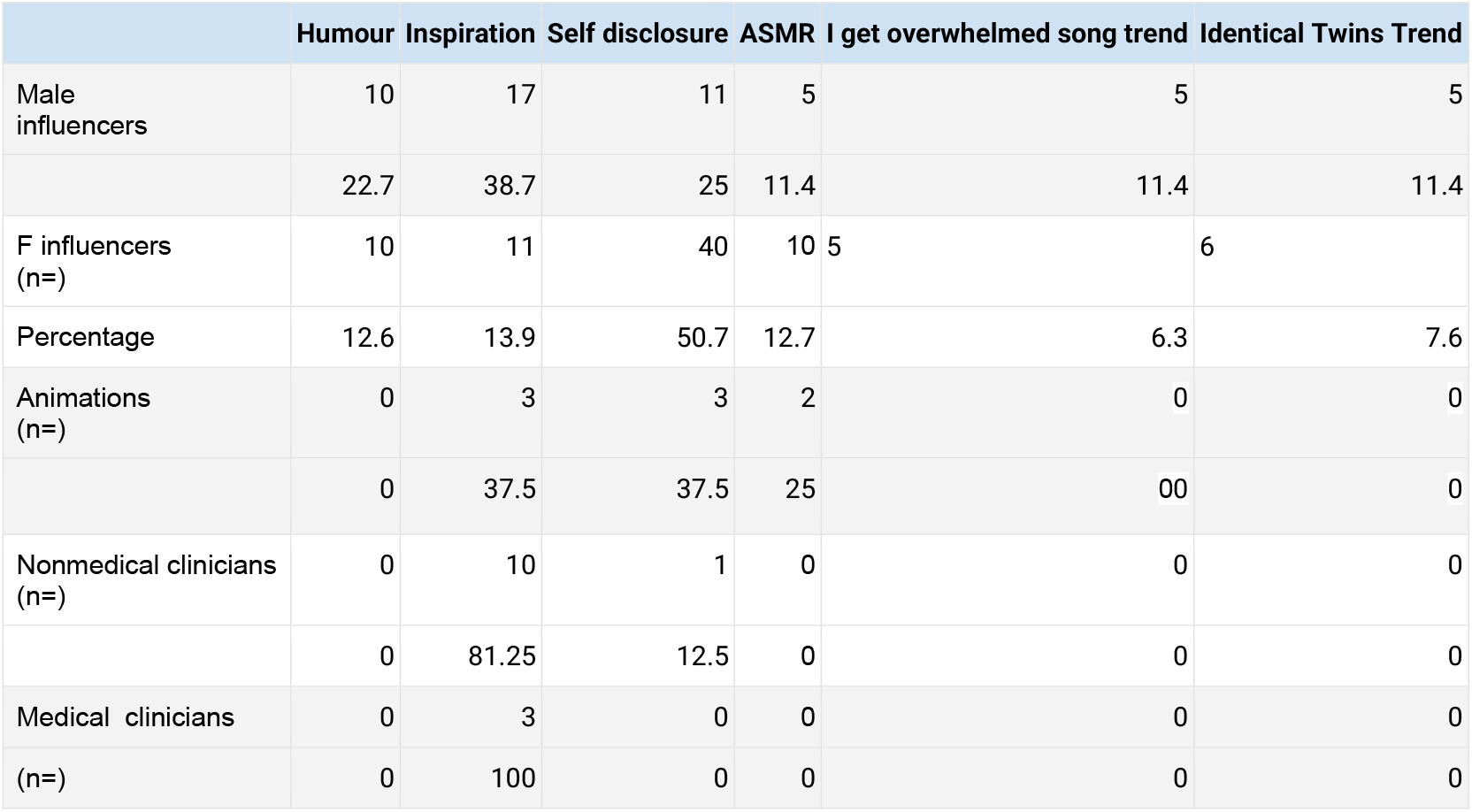
Results - content themes in final data set by creator identity.

### RQ4: What are the characteristics of the data set based on an anxiety management reference checklist?

We coded TikToks in our final data set using a self-care anxiety reference checklist based on the RANZCP Anxiety Guidelines [7]. See table 8. The items in the reference checklist used in this study were Cognitive Behavioral Therapy (CBT) treatment; non-CBT psychological therapies; medication use; education; self-monitoring; positive behaviour; peer engagement; and clinician involvement.

**Table 8:** Results reference data content themes in final data set by creator identity.

| Identity | CBT | Non CBT | Medication use | Education | Self-monitor | Positive behaviour | Peers | Clinician |
| --- | --- | --- | --- | --- | --- | --- | --- | --- |
| M influencers<br>(n=45) | 2 | 2 | 2 | 31 | 36 | 15 | 13 | 4 |
| % | 4.5 | 4.5 | 4.5 | 70.5 | 81.8 | 34.1 | 29.5 | 9.1 |
| F influencers<br>(n=79) | 1 | 6 | 0 | 61 | 63 | 12 | 24 | 1 |
| % | 1.3 | 7.6 | 0 | 77.2 | 79.7 | 15.2 | 30.4 | 1.3 |
| Animations<br>(n=8) | 0 | 0 | 0 | 6 | 7 | 2 | 1 | 0 |
| % | 0 | 0 | 0 | 75 | 87.5 | 25 | 12.5 | 0 |
| Nonmedical clinicians<br>(n=12) | 1 | 3 | 1 | 11 | 9 | 3 | 3 | 7 |
| % | 6.25 | 25 | 6.25 | 81.25 | 68.75 | 18.75 | 25 | 43.75 |
| Physicians<br>(n=3) | 0 | 0 | 0 | 3 | 3 | 0 | 0 | 3 |
| % | 0 | 0 | 0 | 0 | 100 | 0 | 0 | 100 |

Overall we found self-monitoring and positive behaviour themes to be the most frequent in our final data set across all creator identities. We found content themes varied by influencer gender. We found a greater focus on positive behaviours by male influencers (male 34.1%; n=15) versus female influencers (15.2%; n=12). The differences across other creator identities were not notable.

## DISCUSSION

This study aimed to increase the understanding of anxiety content on TikTok and to further develop methods for analysing TikTok video content. We have thus framed the discussion around these two aims.

### Evidence based anxiety self care

We found limited references to evidence-based anxiety self-care content in our final data set. The TikToks in our final data set featured limited discussion of either evidence -based treatment options or how to seek help for anxiety. However, a high proportion of TikToks across all creator identities did feature anxiety education and anxiety self-awareness content. See Table 7. This suggests that the TikToks in our data set were primarily directed at raising awareness of and de-stigmatising anxiety symptoms. We believe that this is a salient finding. Researchers have suggested consumer created content may differ from content produced by clinicians [47]. Adolescents with mental health symptoms demonstrate resource preferences related to symptomatic relief [64]. Conversely, TikToks posted by mental health professionals feature coping techniques, education about anxiety and depression, and how to obtain treatment [50].

We suggest that there may be two reasons for the relative paucity of evidence-based anxiety self-care advice in our final data set. First, we believe this may have been the result of the high proportion of influencer created TikToks in our final data set. Female influencers made up 53.7% (n=79) and male influencers 30.6% (n=45) of our data set. See Table 5. Consumer created content may be especially prevalent in the mental health domain. For example, Devendorf and colleagues similarly reported a high proportion of consumer created YouTube videos about depression [54].

Secondly, the presence of evidence-based self-care content may also reflect content differences between social media platforms. Zheluk et al reported a consistently lower concordance with low back pain guidelines on TikTok when compared with YouTube [30]. TikToks about back pain were consistently simpler, and generally featured single messages. YouTube videos by contrast were longer and more. We believe clinicians and researchers should consider the characteristics of TikTok as a medium, and of the scope of popular anxiety content on TikTok, when seeking to create evidence informed anxiety content.

### Affective content

TikTok anxiety content may be viewed for symptomatic relief beyond evidence-based health information seeking. Researchers have suggested that adolescents with mental health symptoms demonstrate resource preferences related to symptomatic relief [64]. Symptomatic relief may derive from the affective dimension of TikTok content directed at improving emotional self-regulation. TikTok content is likely primarily consumed on smartphones. Smartphone use for emotional regulation has been described in scientific literature. Researchers have described emotional regulation through smartphones as a “buffering effect” [65] or as a distraction strategy [66,67]. Distraction has also been associated with problematic smartphone use [68]. Researchers have also noted females may be more prone to problematic smartphone use [69]. This is salient, as anxiety is also reported to be more common among adolescent females.

ASMR provides an example of TikTok content directed at emotional regulation. ASMR refers to the sensations elicited in response to a range of sounds and visual stimuli [70]. We coded 12 TikToks featuring ASMR in our final data set. These ASMR videos were all from influencers. See Table 7. We believe ASMR provides an example of the use of TikTok for emotional self-regulation. However, the behaviour of TikTok viewing itself may also offer emotional self-regulation. TikTok viewing has been reported to induce physiological changes in subjects in the preoperative environment [67]. In summary, we suggest researchers and clinicians consider the symptomatic relief offered by TikTok use on smartphones by adolescents alongside with the communication of specific evidence-based anxiety messaging.

### Self disclosure

Self-disclosure on TikTok may also provide symptomatic relief to adolescents with anxiety. Consumer created content is the most common category in mental health social media [47,54]. Self-disclosure featured prominently in content created by non-clinicians in our final data set. Self -disclosure featured in 50% of female influencers’ and 25% of male influencers’ TikToks.

Researchers have consistently identified personal narratives and self-disclosure as the most common themes in mental health oriented YouTube [71], TikTok [72] and Facebook [73]. Videos that contain personal narratives and experiential knowledge generate the most user engagement and are preferred sources for users searching for mental health information [71]. Adolescents are more likely to disclose than adults [74]. Online self-disclosure for mental health problems may offer social support acceptance [75], and enhance relationship quality [73] to a degree potentially unavailable to adolescents offline. We suggest that gender and self-disclosure are salient considerations for clinicians seeking to produce highly viewed TikToks about anxiety.

### TikTok trends content

We identified several novel TikTok trends in our final data set. Trends may be music, editing styles, or other features common across TikTok creator identities [57,58]. These trends may be relevant to researchers and clinicians from a user engagement perspective. We identified two trends in our final data set. These trends were the “identical twins” TikTok trend, and the use of the “I get overwhelmed” song trend [60]. We suggest that identification of TikTok trends in a health-related content represents a novel approach to the analysis of TikToks. TikTok trends offer an additional coding category for health professionals seeking to develop engaging mental health content.

## Discussion of methods

We identified consistent differences in content and metadata between aggregated creator identities. Researchers have previously suggested that aggregated creator identities may be useful coding categories with which to analyse the characteristics of specific health problems and social media channels [30,52]. We found male and female influencers were the most common creator identities in our final data set. We observed multiple differences in content created by male and female influencers. Notably, we found self-disclosure was most common among female influencers (female 50%; n =40 versus male 25%; n=11), whereas positive self-behaviour recommendations were most common among male influencers (males 34%; n=15 versus females 15.2%; n=12). We believe that gender, in addition to other identities, should routinely be applied to analysis of TikToks. In summary, through this study we identified novel dimensions to anxiety TikToks, and extended the methods of analysis of TikToks

## LIMITATIONS

We identified several limitations to this study.

TikTok use is growing rapidly. When we gathered the data for this study in December 2021. Global TikTok views for #anxiety were 6 billion at that time. By the time of final writing of this paper in May 2022, the global views for #anxiety had reached 13.9 billion. This rapid growth reflects the dynamism of TikTok as a medium. The rapid growth of TikTok use should also be considered a limitation of the data in this paper. The findings of this paper should thus be considered most relevant to the status of TikTok in late 2021 rather that the date of publication.

We did not seek to demonstrate that adolescents purposefully use TikTok for self-disclosure, emotional regulation, or to find information about anxiety. Rather, we have identified patterns of popular TikToks that featured specific elements of anxiety content. These patterns of content are broadly consistent with earlier research describing mental health content on social media.

We identified a high number of TikToks that were withdrawn between gathering raw data and final analysis (10% of raw data set; n=23) We did not code these videos. We consider this to be a limitation of this study. Much of the influencer created content is very personal, and self-disclosure may be challenging for some creators. Examination of the TikToks withdrawn from public viewing over a specific period may offer researchers and clinicians insights into the scope of sensitive anxiety content available on TikTok.

## FUTURE RESEARCH

We believe that this study suggests several avenues that merit further investigation.

First, we believe the further investigation of TikTok content on mobile phones by adolescents for self-regulation of anxiety merits attention. In particular, the self-directed use of TikToks for managing anxiety symptoms, independent of scientific content. This approach is consistent with the literature describing the use of social media distraction via social media.

Second, believe that longitudinal studies of specific health conditions are warranted. TikTok is a rapidly growing and dynamic medium. The pattern of discussions and creator identities described in this paper reflects the anxiety discussions in December 2021. We believe that repeating this study in 12 months will yield different results and further extend scholarly understanding of this anxiety content on TikTok.

Third, we suggest further research into the thematic categories used to code TikToks. The approach described in this paper suggests coding by content and creator categories such as gender offers novel insights into how anxiety is represented on TikTok. We believe identifying the themes and creator categories in each disease specific data set may represent a valuable direction for future research and targeting messages at specific cohorts.

## CONCLUSIONS

Both social media and anxiety feature prominently in many adolescents’ lives. This study aimed to evaluate the scope of anxiety content available on TikTok in December 2021, and to extend the methods of analysis of anxiety related TikTok content. Our content findings are broadly consistent with previous research into adolescent use of social media. It is plausible that adolescents use TikTok on mobile phones for symptomatic relief and self-care information. This research also provides methodological insights for researchers and clinicians seeking to understand TikTok, and to develop engaging content targeted at the specific concerns and preferences of adolescent TikTok consumers.

## Data Availability

All data produced in the present study are available upon reasonable request to the authors

## REFERENCES

1. Costello EJ, Egger HL, Angold A. The Developmental Epidemiology of Anxiety Disorders: Phenomenology, Prevalence, and Comorbidity. Child Adolesc Psychiatr Clin N Am. 2005;14(4):631–648. doi:10.1016/j.chc.2005.06.003

2. Bandelow B, Michaelis S. Epidemiology of anxiety disorders in the 21st century. Dialogues Clin Neurosci. 2015;17(3):327.

3. Whiteford HA, Degenhardt L, Rehm J, et al. Global burden of disease attributable to mental and substance use disorders: findings from the Global Burden of Disease Study 2010. The Lancet. 2013;382(9904):1575–1586. doi:10.1016/S0140-6736(13)61611-6

4. Sawyer SM, Azzopardi PS, Wickremarathne D, Patton GC. The age of adolescence. Lancet Child Adolesc Health. 2018;2(3):223–228. doi:10.1016/S2352-4642(18)30022-1

5. Kessler RC, Petukhova M, Sampson NA, Zaslavsky AM, Wittchen HU. Twelve-month and lifetime prevalence and lifetime morbid risk of anxiety and mood disorders in the United States. Int J Methods Psychiatr Res. 2012;21(3):169–184.

6. Kessler RC, Avenevoli S, Costello EJ, et al. Prevalence, persistence, and sociodemographic correlates of DSM-IV disorders in the National Comorbidity Survey Replication Adolescent Supplement. Arch Gen Psychiatry. 2012;69(4):372–380.

7. Andrews G, Bell C, Boyce P, et al. Royal Australian and New Zealand College of Psychiatrists clinical practice guidelines for the treatment of panic disorder, social anxiety disorder and generalised anxiety disorder. Aust N Z J Psychiatry. 2018;52(12):1109–1172.

8. Anxiety disorders - Symptoms and causes. Mayo Clinic. Accessed September 11, 2022. https://www.mayoclinic.org/diseases-conditions/anxiety/symptoms-causes/syc-20350961

9. Bandelow B, Reitt M, Röver C, Michaelis S, Görlich Y, Wedekind D. Efficacy of treatments for anxiety disorders: a meta-analysis. Int Clin Psychopharmacol. 2015;30(4):183–192. doi:10.1097/YIC.0000000000000078

10. Sartorius N, Ustün TB, Lecrubier Y, Wittchen HU. Depression comorbid with anxiety: results from the WHO study on psychological disorders in primary health care. Br J Psychiatry Suppl. 1996;(30):38–43.

11. Alonso J, Lépine JP, ESEMeD/MHEDEA 2000 Scientific Committee. Overview of key data from the European Study of the Epidemiology of Mental Disorders (ESEMeD). J Clin Psychiatry. 2007;68 Suppl 2:3–9.

12. Aichner T, Grünfelder M, Maurer O, Jegeni D. Twenty-Five Years of Social Media: A Review of Social Media Applications and Definitions from 1994 to 2019. Cyberpsychology Behav Soc Netw. 2021;24(4):215–222. doi:10.1089/cyber.2020.0134

13. Auxier B, MONICA, ERSON. Social Media Use in 2021. Pew Research Center: Internet, Science & Tech. Published April 7, 2021. Accessed October 30, 2021. https://www.pewresearch.org/internet/2021/04/07/social-media-use-in-2021/

14. Koetsier J. Top 10 Most Downloaded Apps And Games Of 2021: TikTok, Telegram Big Winners. Forbes. Accessed September 11, 2022. https://www.forbes.com/sites/johnkoetsier/2021/12/27/top-10-most-downloaded-apps-and-games-of-2021-tiktok-telegram-big-winners/

15. bbc.co.uk. TikTok overtakes YouTube for average watch time in US and UK. BBC News. https://www.bbc.com/news/technology-58464745. Published September 6, 2021. Accessed October 31, 2021.

16. Literat I. “Teachers Act Like We’re Robots”: TikTok as a Window Into Youth Experiences of Online Learning During COVID-19. AERA Open. 2021;7:2332858421995537. doi:10.1177/2332858421995537

17. Holmes R. Is COVID-19 Social Media’s Levelling Up Moment? Forbes. Accessed September 11, 2022. https://www.forbes.com/sites/ryanholmes/2020/04/24/is-covid-19-social-medias-levelling-up-moment/

18. Kids now spend nearly as much time watching TikTok as YouTube in US, UK and Spain TechCrunch. Accessed September 11, 2022. https://techcrunch.com/2020/06/04/kids-now-spend-nearly-as-much-time-watching-tiktok-as-youtube-in-u-s-u-k-and-spain/

19. Teens, Social Media and Technology 2022 Pew Research Center. Accessed August 15, 2022. https://www.pewresearch.org/internet/2022/08/10/teens-social-media-and-technology-2022/

20. Smith A, Anderson M. Social Media Use in 2018. Pew Research Center: Internet, Science & Tech. Published March 1, 2018. Accessed September 27, 2022. https://www.pewresearch.org/internet/2018/03/01/social-media-use-in-2018/

21. Wang N, Sun Y. Social influence or personal preference? Examining the determinants of usage intention across social media with different sociability. Inf Dev. 2016;32(5):1442–1456. doi:10.1177/0266666915603224

22. Chang CM, Hsu MH. Understanding the determinants of users’ subjective well-being in social networking sites: an integration of social capital theory and social presence theory. Behav Inf Technol. 2016;35(9):720–729. doi:10.1080/0144929X.2016.1141321

23. Whiting A, Williams D. Why people use social media: a uses and gratifications approach. Qual Mark Res Int J. 2013;16(4):362–369. doi:10.1108/QMR-06-2013-0041

24. Bossen CB, Kottasz R. Uses and gratifications sought by pre-adolescent and adolescent TikTok consumers. Young Consum. Published online 2020.

25. Wang J. From Banning to Regulating TikTok: Addressing Concerns of National Security, Privacy, and Online Harms. The Foundation Law Justice and Society; 2020. https://www.fljs.org/sites/default/files/migrated/publications/From%20Banning%20to%20Regulating%20TikTok.pdf

26. Montag C, Yang H, Elhai JD. On the Psychology of TikTok Use: A First Glimpse From Empirical Findings. Front Public Health. 2021;9:641673. doi:10.3389/fpubh.2021.641673

27. Moreno MA, Whitehill JM. Influence of Social Media on Alcohol Use in Adolescents and Young Adults. Alcohol Res Curr Rev. 2014;36(1):91–100.

28. Guessoum SB, Lachal J, Radjack R, et al. Adolescent psychiatric disorders during the COVID-19 pandemic and lockdown. Psychiatry Res. 2020;291:113264. doi:10.1016/j.psychres.2020.113264

29. Zenone M, Ow N, Barbic S. TikTok and public health: a proposed research agenda. BMJ Glob Health. 2021;6(11):e007648.

30. Zheluk A, Anderson J, Dineen-Griffin S. Analysis of Acute Non-specific Back Pain Content on TikTok: An Exploratory Study. Cureus. 2022;14(1). doi:10.7759/cureus.21404

31. Sha P, Dong X. Research on Adolescents Regarding the Indirect Effect of Depression, Anxiety, and Stress between TikTok Use Disorder and Memory Loss. Int J Environ Res Public Health. 2021;18(16):8820. doi:10.3390/ijerph18168820

32. Gao J, Zheng P, Jia Y, et al. Mental health problems and social media exposure during COVID-19 outbreak. PLOS ONE. 2020;15(4):e0231924. doi:10.1371/journal.pone.0231924

33. Vannucci A, Flannery KM, Ohannessian CM. Social media use and anxiety in emerging adults. J Affect Disord. 2017;207:163–166. doi:10.1016/j.jad.2016.08.040

34. Andreassen HK, Bujnowska-Fedak MM, Chronaki CE, et al. European citizens’ use of E-health services: a study of seven countries. BMC Public Health. 2007;7(1):53.

35. Keles B, McCrae N, Grealish A. A systematic review: the influence of social media on depression, anxiety and psychological distress in adolescents. Int J Adolesc Youth. 2020;25(1):79–93. doi:10.1080/02673843.2019.1590851

36. Nesi J. The impact of social media on youth mental health: challenges and opportunities. N C Med J. 2020;81(2):116–121.

37. Galea S, Merchant RM, Lurie N. The Mental Health Consequences of COVID-19 and Physical Distancing: The Need for Prevention and Early Intervention. JAMA Intern Med. 2020;180(6):817–818. doi:10.1001/jamainternmed.2020.1562

38. da Silva ML, Rocha RSB, Buheji M, Jahrami H, Cunha K da C. A systematic review of the prevalence of anxiety symptoms during coronavirus epidemics. J Health Psychol. 2021;26(1):115–125. doi:10.1177/1359105320951620

39. Douglas M, Katikireddi SV, Taulbut M, McKee M, McCartney G. Mitigating the wider health effects of covid-19 pandemic response. BMJ. 2020;369:m1557. doi:10.1136/bmj.m1557

40. Matias T, Dominski FH, Marks DF. Human needs in COVID-19 isolation. J Health Psychol. 2020;25(7):871–882. doi:10.1177/1359105320925149

41. de Figueiredo CS, Sandre PC, Portugal LCL, et al. COVID-19 pandemic impact on children and adolescents’ mental health: Biological, environmental, and social factors. Prog Neuropsychopharmacol Biol Psychiatry. 2021;106:110171. doi:10.1016/j.pnpbp.2020.110171

42. Harwood E. TikTok, identity struggles, and mental health issues: how are the youth of today coping? In: Curtin University; 2021. http://www.networkconference.netstudies.org/2021/wp-content/uploads/2021/04/TikTok-identity-struggles-and-mental-health-issues-How-are-the-youth-of-today-coping.pdf

43. Comp G, Dyer S, Gottlieb M. Is TikTok The Next Social Media Frontier for Medicine? AEM Educ Train. 2020;5(3):10.1002/aet2.10532. doi:10.1002/aet2.10532

44. Singh S, Roy D, Sinha K, Parveen S, Sharma G, Joshi G. Impact of COVID-19 and lockdown on mental health of children and adolescents: A narrative review with recommendations. Psychiatry Res. 2020;293:113429. doi:10.1016/j.psychres.2020.113429

45. Zhong B, Huang Y, Liu Q. Mental health toll from the coronavirus: Social media usage reveals Wuhan residents’ depression and secondary trauma in the COVID-19 outbreak. Comput Hum Behav. 2021;114:106524. doi:10.1016/j.chb.2020.106524

46. Zhao N, Zhou G. Social Media Use and Mental Health during the COVID-19 Pandemic: Moderator Role of Disaster Stressor and Mediator Role of Negative Affect. Appl Psychol Health Well-Being. 2020;12(4):1019–1038. doi:10.1111/aphw.12226

47. MacLean SA, Basch CH, Reeves R, Basch CE. Portrayal of generalized anxiety disorder in YouTube™ videos. Int J Soc Psychiatry. 2017;63(8):792–795. doi:10.1177/0020764017728967

48. Zhang B, Zaman A, Silenzio V, Kautz H, Hoque E. The Relationships of Deteriorating Depression and Anxiety With Longitudinal Behavioral Changes in Google and YouTube Use During COVID-19: Observational Study. JMIR Ment Health. 2020;7(11):e24012. doi:10.2196/24012

49. Eghtesadi M, Florea A. Facebook, Instagram, Reddit and TikTok: a proposal for health authorities to integrate popular social media platforms in contingency planning amid a global pandemic outbreak. Can J Public Health Rev Can Santé Publique. 2020;111(3):389–391. doi:10.17269/s41997-020-00343-0

50. Sood A. TikTok: A new playground for the child psychiatrist. J Am Acad Child Adolesc Psychiatry. Published online 2021:S5–S5.

51. Azer SA. Are DISCERN and JAMA Suitable Instruments for Assessing YouTube Videos on Thyroid Cancer? Methodological Concerns. J Cancer Educ Off J Am Assoc Cancer Educ. Published online May 29, 2020. doi:10.1007/s13187-020-01763-9

52. Zheluk A, Maddock J. Plausibility of Using a Checklist With YouTube to Facilitate the Discovery of Acute Low Back Pain Self-Management Content: Exploratory Study. JMIR Form Res. 2020;4(11):e23366. doi:10.2196/23366

53. Nord A. TikTok Scraper & Downloader. Published online 2021. Accessed October 30, 2021. https://www.npmjs.com/package/tiktok-scraper

54. Devendorf A, Bender A, Rottenberg J. Depression presentations, stigma, and mental health literacy: A critical review and YouTube content analysis. Clin Psychol Rev. 2020;78:101843. doi:10.1016/j.cpr.2020.101843

55. Thomas DR. A General Inductive Approach for Analyzing Qualitative Evaluation Data. Am J Eval. 2006;27(2):237–246. doi:10.1177/1098214005283748

56. Guo JZ, Chong KPL, Woo BKP. Utilizing YouTube as platform for psychiatric emergency patient outreach in Chinese Americans. Asian J Psychiatry. 2020;50:101960. doi:10.1016/j.ajp.2020.101960

57. Carey E. This Trending TikTok Sound Is An Easy Way To Roast Anyone You Want. Bustle. Accessed June 6, 2022. https://www.bustle.com/life/me-talking-to-tiktok-trend

58. Jaudon E. TikTok trends: How to find them and make them your own. Bazaarvoice. Published March 18, 2022. Accessed June 6, 2022. https://www.bazaarvoice.com/blog/tiktok-trends-how-to/

59. Connor DeWolfe on TikTok. TikTok. Accessed September 11, 2022. https://www.tiktok.com/@connordewolfe/video/6997467882389064966

60. Royal & the Serpent - Overwhelmed (Official Music Video).; 2020. Accessed June 6, 2022. https://www.youtube.com/watch?v=_e7UYTY96Xs

61. Sampson M, Cumber J, Li C, Pound CM, Fuller A, Harrison D. A systematic review of methods for studying consumer health YouTube videos, with implications for systematic reviews. PeerJ. 2013;1:e147. doi:10.7717/peerj.147

62. Drozd B, Couvillon E, Suarez A. Medical YouTube Videos and Methods of Evaluation: Literature Review. JMIR Med Educ. 2018;4(1):e3. doi:10.2196/mededu.8527

63. Cascio MA, Lee E, Vaudrin N, Freedman DA. A team-based approach to open coding: Considerations for creating intercoder consensus. Field Methods. 2019;31(2):116–130.

64. Toscos T, Coupe A, Flanagan M, et al. Teens Using Screens for Help: Impact of Suicidal Ideation, Anxiety, and Depression Levels on Youth Preferences for Telemental Health Resources. JMIR Ment Health. 2019;6(6):e13230. doi:10.2196/13230

65. Marzouki Y, Aldossari FS, Veltri GA. Understanding the buffering effect of social media use on anxiety during the COVID-19 pandemic lockdown. Humanit Soc Sci Commun. 2021;8(1):1–10.

66. Throuvala MA, Pontes HM, Tsaousis I, Griffiths MD, Rennoldson M, Kuss DJ. Exploring the Dimensions of Smartphone Distraction: Development, Validation, Measurement Invariance, and Latent Mean Differences of the Smartphone Distraction Scale (SDS). Front Psychiatry. 2021;12. Accessed June 6, 2022. https://www.frontiersin.org/article/10.3389/fpsyt.2021.642634

67. Gu S, Ping J, Xu M, Zhou Y. TikTok browsing for anxiety relief in the preoperative period: A randomized clinical trial. Complement Ther Med. 2021;60:102749.

68. Throuvala MA, Griffiths MD, Rennoldson M, Kuss DJ. Mind over Matter: Testing the Efficacy of an Online Randomized Controlled Trial to Reduce Distraction from Smartphone Use. Int J Environ Res Public Health. 2020;17(13):4842. doi:10.3390/ijerph17134842

69. van Oosten Jmf, Vandenbosch L. Sexy online self-presentation on social network sites and the willingness to engage in sexting: A comparison of gender and age. J Adolesc. 2017;54:42–50. doi:10.1016/j.adolescence.2016.11.006

70. Poerio GL, Blakey E, Hostler TJ, Veltri T. More than a feeling: Autonomous sensory meridian response (ASMR) is characterized by reliable changes in affect and physiology. PLOS ONE. 2018;13(6):e0196645. doi:10.1371/journal.pone.0196645

71. Oliphant T. User Engagement with Mental Health Videos on YouTube. J Can Health Libr Assoc J Assoc Bibl Santé Can. 2013;34(3):153–158. doi:10.5596/c13-057

72. Nabity-Grover T, Cheung CMK, Thatcher JB. Inside out and outside in: How the COVID-19 pandemic affects self-disclosure on social media. Int J Inf Manag. 2020;55:102188. doi:10.1016/j.ijinfomgt.2020.102188

73. Lerman BI, Lewis SP, Lumley M, Grogan GJ, Hudson CC, Johnson E. Teen Depression Groups on Facebook: A Content Analysis. J Adolesc Res. 2017;32(6):719–741. doi:10.1177/0743558416673717

74. Nosko A, Wood E, Molema S. All about me: Disclosure in online social networking profiles: The case of FACEBOOK. Comput Hum Behav. 2010;26(3):406–418. doi:10.1016/j.chb.2009.11.012

75. Schlosser AE. Self-disclosure versus self-presentation on social media. Curr Opin Psychol. 2020;31:1–6. doi:10.1016/j.copsyc.2019.06.025

